# High temperature has no impact on the reproduction number and new cases of COVID-19 in Bushehr, Iran

**DOI:** 10.1101/2020.06.14.20130906

**Authors:** Ebrahim Sahafizadeh, Samaneh Sartoli

## Abstract

**Background:** COVID-19 was first reported in Iran on February 19, 2020. Bushehr, one of the warmest provinces of Iran, was the last province confirmed to be infected on March 5, 2020. In the beginning of April, Bushehr was announced as a ‘white’, coronavirus-free, province. However, increasing the temperature in the next months did not affect the spread of coronavirus and the number of confirmed cases increased during the next months, so that Bushehr was announced as a ‘Red’ province on June 13, 2020.

**Methods:** This paper aims 1) to estimate the reproduction number of COVID-19 in Bushehr considering COVID-19 reported cases of Bushehr from April to June 12, 2020, using exponential function and SIR epidemic model, and 2) to investigate the impact of temperature on the reproduction number and the spread of coronavirus in Bushehr using the temperature data.

**Results:** The reproduction number was estimated to be 2.564, 2.641 and 2.573 at the beginning of April, May and June 2020 respectively. Regarding the increase in the temperature from April to June, the results showed that not only was the spread of COVID-19 not reduced but it also increased.

**Conclusion:** Data analysis on this study showed that high temperature has no impact on the reproduction number and does not slow down the spread of coronavirus in Bushehr.

## Background

On December 31, 2019, the novel coronavirus (COVID-19) emerged in Wuhan, China, and rapidly spread globally [1]. The first confirmed cases of COVID-19 in Iran were reported on February 19, 2020 in Qom [2]. Although the outbreak quickly moved to other provinces of Iran and the basic reproduction number was estimated to be more than four [3], Bushehr was the last province confirmed to be infected in the third week of the outbreak (on March 5, 2020).

Bushehr is located in south west of Iran with a long coastline onto the Persian Gulf. Bushehr is one of the warmest provinces of Iran and attracts many tourists in winter and Nowruz (Persian New Year) holidays every year. In this case, according to the idea that high temperature and humidity slow down the spread of coronavirus, after the outbreak in Iran, many people rushed to Bushehr and other provinces in south of Iran. This phenomenon led to a strong travelling ban in this province and people spontaneously quarantined themselves. As the other provinces of Iran, schools, universities, mosques, Friday prayers and social events closed in Bushehr and ‘stay at home’ policy was implemented. Hence, till the end of March 2020 cumulative number of reported cases remained below 100. On April 5, 2020, the president announced Bushehr as the white province and authorized the local authorities to reopen the schools and universities. On May 3, 2020, five counties of Bushehr were announced as white counties.

The social distancing measures kept the number of new cases down in Bushehr. Reporting Bushehr as a white region, opening many businesses, increasing contact rate, stop using face masks as the temperature increased and ignoring social distancing measures increased the number of COVID-19 confirmed cases in Bushehr, so that it was announced as a ‘Red’ province on June 13, 2020. The aim of this study is to estimate the reproduction number of COVID-19 and investigate the effect of temperature on the reproduction number and the spread of COVID-19 in Bushehr.

## Methods

We used the temperature data and COVID-19 reported cases of Bushehr from April to June 12, 2020, reported by Bushehr meteorological organization [4] and Bushehr Province University of Medical Science [5] respectively to investigate the impact of temperature on the spread of COVID-19 in Bushehr. Fig. 1 shows the number of cumulative cases and Fig. 2 shows the daily new confirmed cases in Bushehr.

**Fig 1.**
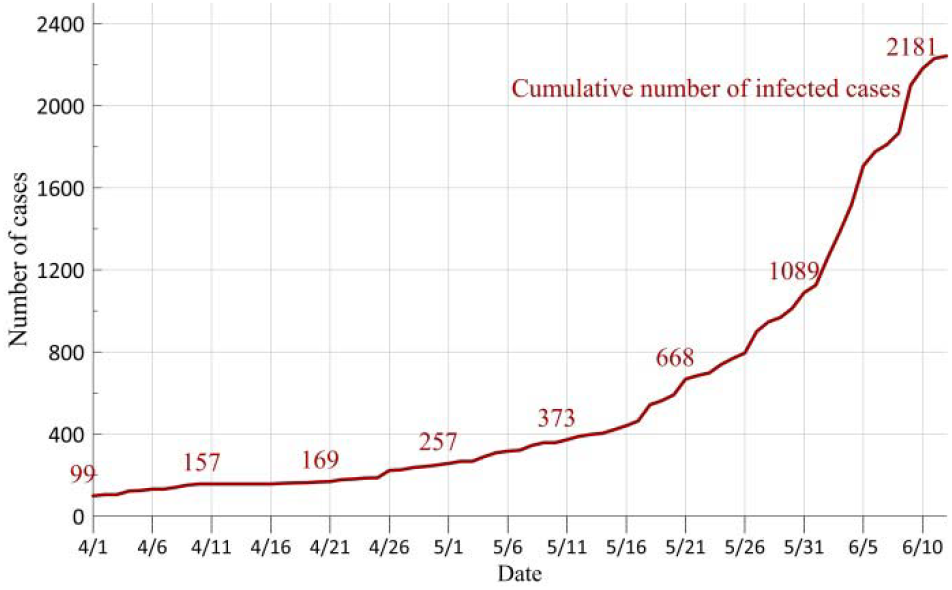
Cumulative infected cases of Bushehr

**Fig 2.**
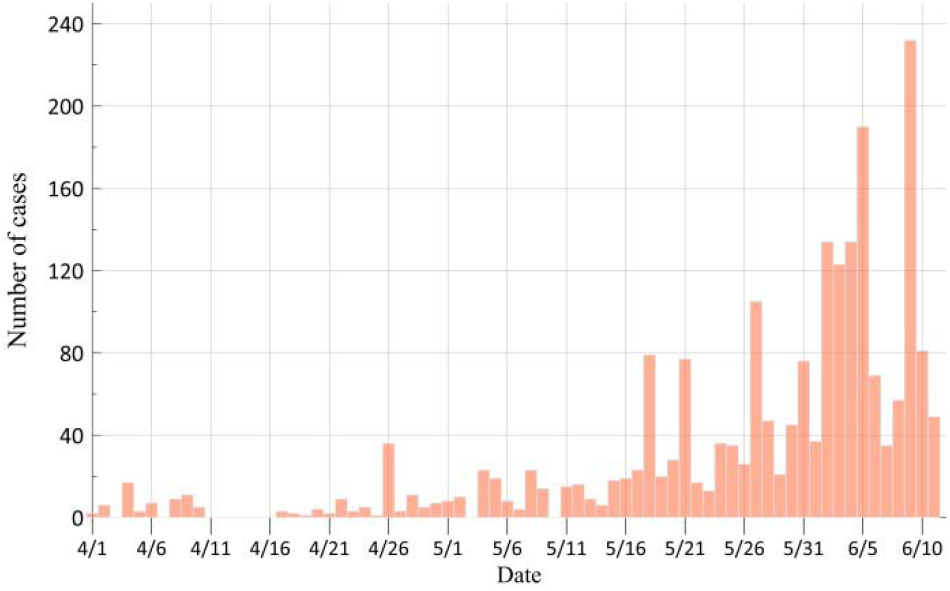
Daily new confirmed cases of Bushehr

We employ SIR (Susceptible-Infected-Removed) epidemic modelling [6] to model the spreading process of COVID-19 in Bushehr, and exponential growth rate [7] to estimate the parameters and effective reproduction number. Runge-Kutta method is used to solve the ordinary differential equations and the simulations are conducted in MATLAB. We use Root Mean Squared Error (RMSE) to represent the spatial error in order to show the goodness of fit.

## Results

We used exponential function to fit the number of infected cases. The exponential function is a widely used function to predict the trend of epidemics as follows.

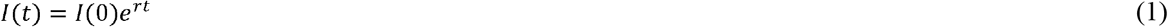

Where *I*(*t*) is the number of infected cases at time t and J(0) is the number of infected cases at time 0.In this case 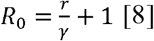, where R is the basic reproduction number and is the remove rate. We set *I*(0) = 12 as the number of infected cases on March 31, 2020 and estimate the parameter r as the growth rate in order to fit exponential function to the reported data. Fig. 3 shows the plot of infected cases and exponential function to fit the infected cases. The x-axis shows the number of days from April 1, 2020 (March 31, 2020 is considered as the origin) and y-axis shows the number of infected cases. The exponential function conforming infected cases was estimated to be J(t) = 12 X e^0.06551^ in which *I*(0) = 12 is the number of infected cases on March 31, 2020. The coefficient 0.06551 is estimated with 95% confidence bound. As shown in the Fig. 3, RMSE is 9.391 and R-square is 0.9994. The estimated reproduction number is 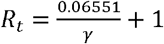 where γ is the remove rate which is the inverse of infectious period. To estimate y we use SIR model as follows.

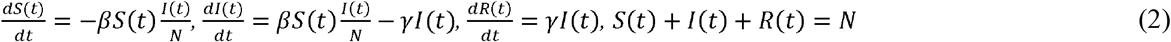

Where *S*(*t*), *I*(*t*) and *R*(*t*) are the number of susceptible, infected and removed people and N is assumed to be the population of Bushehr province 1163400 in 2016 [9]. We set γ = β − 0.06551 and fit SIR curve (Fig. 4) to reported data with the least RMSE and γ is estimated to be 0.04188, Hence,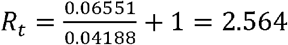.

**Fig 3.**
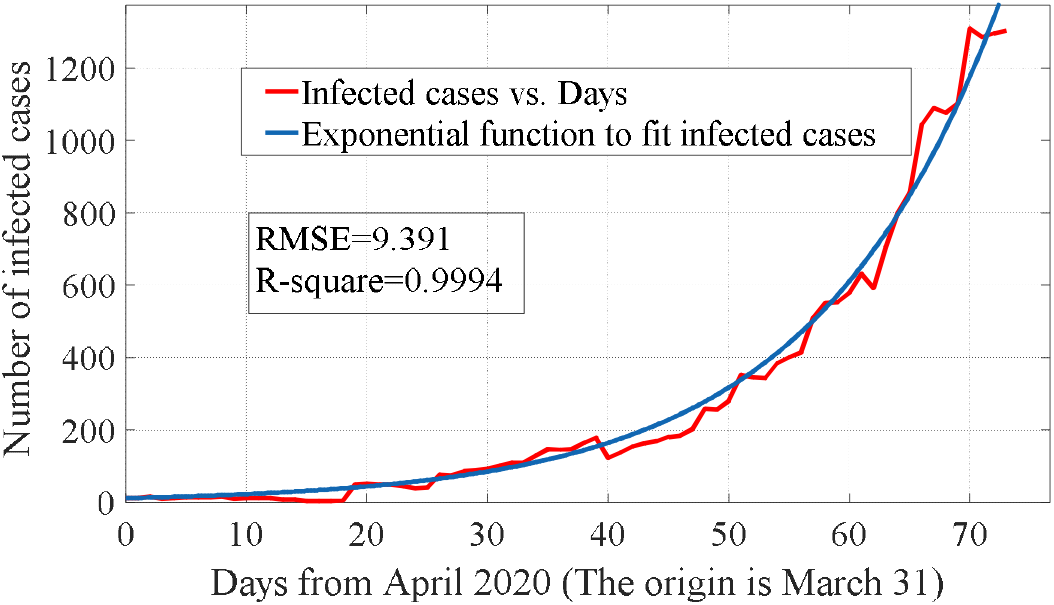
The exponential function to fit the number of infected cases

**Fig 4.**
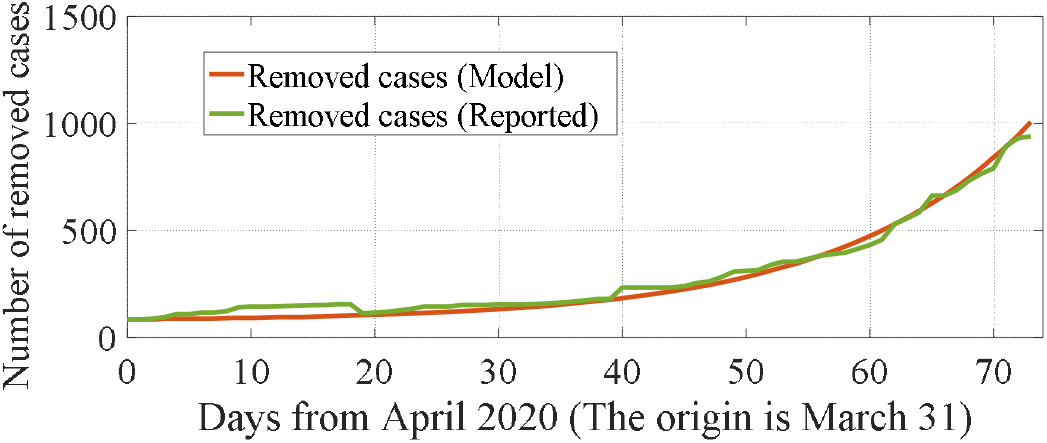
The SIR curve to fit the number of removed cases

We repeated the same process and estimated *R*_*t*_ for different time periods. Table 1. shows the estimated parameters and reproduction number at the first day of April, May and June 2020.

**Table 1.**
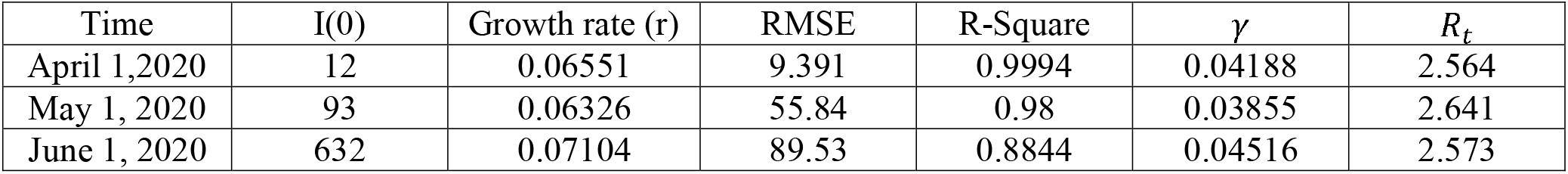
Estimated parameters and reproduction number of COVID-19 in Bushehr in different time periods.

| Time | I(0) | Growth rate (r) | RMSE | R-Square | $\gamma$ | $R_t$ |
| --- | --- | --- | --- | --- | --- | --- |
| April 1,2020 | 12 | 0.06551 | 9.391 | 0.9994 | 0.04188 | 2.564 |
| May 1, 2020 | 93 | 0.06326 | 55.84 | 0.98 | 0.03855 | 2.641 |
| June 1, 2020 | 632 | 0.07104 | 89.53 | 0.8844 | 0.04516 | 2.573 |

## Discussion

As it was mentioned, Bushehr is one of the warmest provinces in Iran with high temperature and air humidity. From April, the temperature increased and air conditioners were turned on everywhere, but in contrast to the findings of previous studies [10][11] which indicate that high temperature reduces the spread of coronavirus, the number of infected cases increased. We used the temperature data from April and June 12, 2020 to investigate the effect of temperature on the spread of COVID-19 in Bushehr. Fig. 5 shows the daily temperature and Table 2. shows the monthly average maximum temperature of Bushehr province. As can be seen in Table 2., the average maximum temperature of Bushehr province is 31.72 °C in April, 37.41 °C in May and 41.67 in June, 2020.

**Table 2.** Monthly average maximum temperature of Bushehr province

| Month | Average Maximum Temperature | Standard Deviation |
| --- | --- | --- |
| April, 2020 | 31.72 °C | 3.06 |
| May, 2020 | 37.41 °C | 1.83 |
| June, 2020 | 41.67 °C | 1.96 |

**Fig 5.**
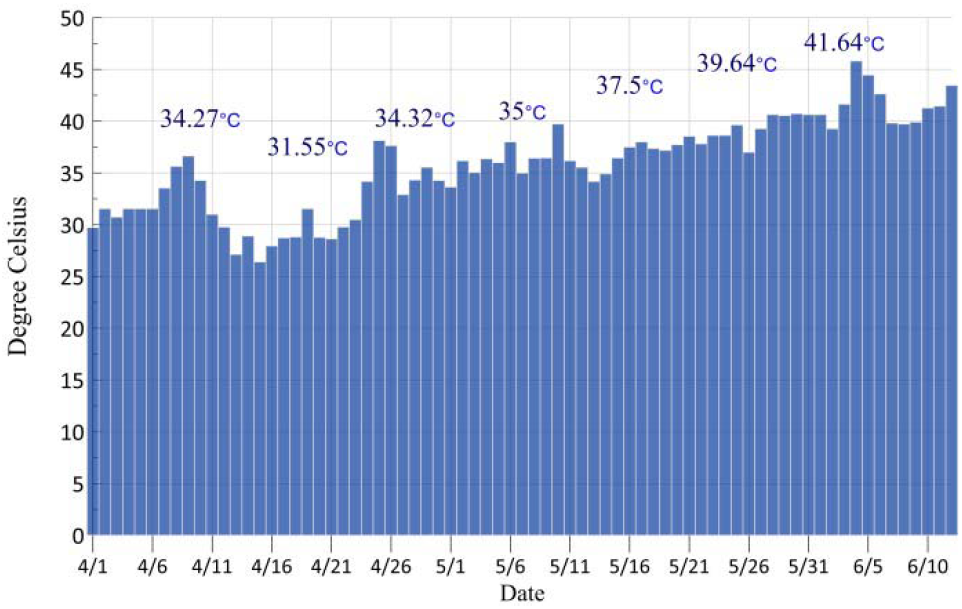
The daily maximum temperature of Bushehr province

Fig. 6 shows the new confirmed cases in April, May and June (until June 12, 2020). Although the temperature in May is significantly higher than April in Bushehr province; the number of new cases in May is four times greater than that in April. Moreover the number of new cases until June 12, 2020, is more than the new cases of May; however the temperature of June is higher than May. It can be concluded that high temperature does not slow down the coronavirus spreading and social distancing implementation is required to control the spread of COVID-19. Moreover, according to Table 1. It can be seen that the increasing temperature does not reduce the reproduction number of COVID-19 in Bushehr.

**Fig 6.**
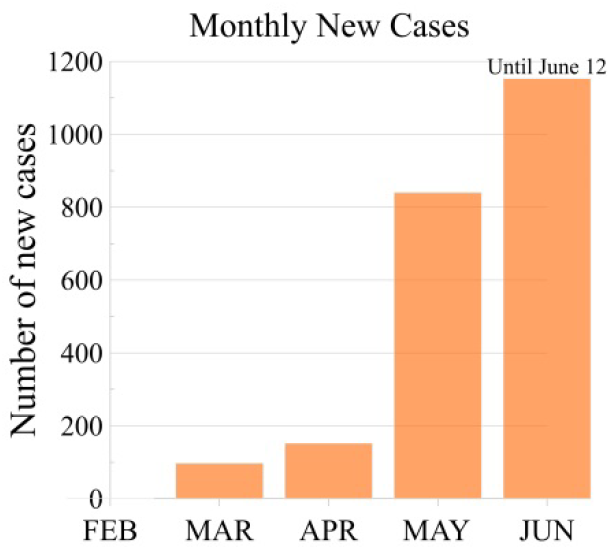
Monthly new confirmed cases

## Conclusion

Bushehr was the last province to be infected by COVID-19 in Iran. Due to the strict social distancing implementation, the cumulative number of infected cases was below 100 by the end of March and Bushehr was announced as a ‘white’, coronavirus-free, province in the beginning of April. Increasing the temperature and the idea that high temperature reduces the spread of coronavirus, announcing Bushehr as a ‘white’ province, social distancing reduction, avoiding the use of face mask and increasing contact rate led to increase the number of COVID-19 cases in Bushehr, such that at June 13, 2020 Bushehr was announced as a ‘Red’ province.

In this paper we estimated the reproduction number of COVID-19 in Bushehr and investigated the effect of high temperature on the reproduction number and new cases of COVID-19 in Bushehr. The reproduction number was estimated to be 2.564, 2.641 and 2.573 at the beginning of April, May and June 2020 respectively. We showed that the increasing temperature has no impact on the reproduction number and the spread of COVID-19 in Bushehr. The reproduction number is significantly higher than one and COVID-19 can poses a tremendous burden on Bushehr’s health care system and society. Social distancing, active case detection, contact tracing and quarantining of contacts will need to be maintained until an effective vaccine becomes available.

## Data Availability

The data that support the findings of this study are available from the corresponding author, upon reasonable request.

## Abbreviations

SARS-CoV-2; nonpharmaceutical interventions; isolation; quarantine; mitigation; public health emergency of international concern; pandemic

## Authors’ contributions

E.S. analyzed the data and wrote the manuscript; S.S. collected data and revised and edited the manuscript. All authors read and approved the final manuscript.

## Ethics approval and consent to participate

Not applicable.

## Consent for publication

Not applicable.

